# Early epidemiological analysis of the 2019-nCoV outbreak based on a crowdsourced data

**DOI:** 10.1101/2020.01.31.20019935

**Authors:** Kaiyuan Sun, Jenny Chen, Cécile Viboud

## Abstract

As the outbreak of novel 2019 coronavirus (2019-nCoV) progresses within China and beyond, there is a need for rapidly available epidemiological data to guide situational awareness and intervention strategies. Here we present an effort to compile epidemiological information on 2019-nCoV from media news reports and a physician community website (dxy.cn) between Jan 20, 2020 and Jan 30, 2020, as the outbreak entered its 7^th^ week. We compiled a line list of patients reported in China and internationally and daily case counts by Chinese province. We describe the demographics, hospitalization and reporting delays for 288 patients, over time and geographically. We find a decrease in case detection lags in provinces outside of Wuhan and internationally, compared to Wuhan, and after Jan 18, 2020, as outbreak awareness increased. The rapid progression of reported cases in different provinces of China is consistent with local transmission beyond Wuhan. The age profile of cases points at a deficit among children under 15 years of age, possibly related to prior immunity with related coronavirus or behavioral differences. Overall, our datasets, which have been publicly available since Jan 21, 2020, align with official reports from Chinese authorities published more than a week later. Availability of publicly available datasets in the early stages of an outbreak is important to encourage disease modeling efforts by independent academic modeling teams and provide robust evidence to guide interventions.

## Introduction

As the outbreak of novel 2019 coronavirus (2019-nCoV) is rapidly expanding in China and beyond, threatening to become a worldwide pandemic ^1^, there is a need for real-time analyses of epidemiological data to increase situational awareness and inform interventions ^2^. In the past, real-time analyses have shed light on the transmissibility, severity, and natural history of an emerging pathogen in the first few weeks of an outbreak ^3-6^. Analyses of detailed line lists of patients are particularly useful to infer key epidemiologic parameters, such as the incubation and infectious periods, and lags in detection, isolation, and reporting of cases ^3,4^. However, individual patient data rarely become publicly available in the first weeks of an outbreak, when the information is most needed.

Building on prior experience collating news reports to monitor Ebola transmission ^7^, here we present an effort to compile individual patient information and subnational epidemic curves on 2019-nCoV from a variety of online resources. Data were made publicly available in real-time and were used by the infectious disease modeling community to generate and compare epidemiological estimates relevant for interventions. Here we describe the process to compile data and provide an early analysis of the demographics, hospitalization, and reporting lags, in different provinces of China and internationally as of Jan 30, 2020.

## Method

### 2019-nCoV data compilation

We used crowdsourced reports from <u>DXY.cn</u>, a Chinese online community network for physicians, health care professionals, pharmacies and healthcare facilities established in 2000 ^8^. An online platform provides real-time coverage of the 2019-nCoV outbreak in China, obtained by collating and curating reports from news media, government TV, national and provincial health agencies. The information reported includes time-stamped cumulative case counts of 2019-nCoV, outbreak maps, and real-time streaming of health authority announcements in Chinese (directly or through state media) ^9^. Every report is linked to an on-line source, which can be followed through for more detailed information on individual cases.

We closely monitored updates on DXY.cn between Jan 20, 2020 and Jan 30, 2020 to extract key information on individual patients in near real-time, as well as daily reported case counts. For individual-level patient information, we used descriptions from the original source in Chinese to retrieve age, gender, province of identification, travel history, reporting date, date of symptom onset and hospitalization, and discharge status, when available. Individual level data were formatted into a line-list database for further quantitative analysis. Individual-level patient data were entered from DXY.cn by a native Chinese speaker (KS), who also generated an English summary for each case. Entries were doubled checked by a second person (JC). Since DXY.cn primarily provides information on cases reported in China, we also compiled additional information on internationally exported cases from global news media.

Further, we reconstructed the daily progression of reported cases in each province of China from Jan 18, 2020 to Jan 30, 2020. We used the daily outbreak situation reports communicated by provincial health authorities, covered by state TV and media, and posted on DXY.cn. All cases in our databases are laboratory confirmed 2019-nCoV patients.

The database was made available as a Google sheet disseminated via Twitter on Jan 21, 2020 and posted on a university website on Jan 24, 2020 ^10^.

### Statistical Analysis

We assessed the age distribution of 2019-nCoV cases by discharge status, adjusted for the demographics of the Chinese population. We used 2016 population estimates from the Institute for Health Metrics and Evaluation ^11^ to calculate the relative risk (RR) of 2019-nCoV infection by age group, following Lemaitre et al ^12^. The RR for age group *i* is defined as:

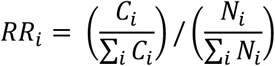

Where *C*_*i*_ is the number of cases in group *i* and *N*_*i*_ is the population of age group *i*.

To estimate trends in the strength of case detection and interventions, we analyzed delays between symptom onset and visit to a healthcare provider, at a hospital or clinic, and from hospitalization to reporting, by time period and location. We considered the period before and after Jan 18, 2020, when media attention and awareness of the outbreak became more pronounced ^13^. Further, intense travel restrictions in and out of the Wuhan region were put in place starting Jan 22, 2020 ^13^.

## Results

### Patients characteristics

Our line list comprised 288 cases reported between Jan 20, 2020 and Jan 29, 2020 including 200 cases from mainland China and 88 international cases (Table 1). The gender ratio was skewed towards male (63%). Within mainland China, there was a large representation of 5 of the 30 provinces, with 17% of cases reported from Hubei (capital, Wuhan), 13% from Shanxi, 8.8% from Tianjin, and 7% each from Yunnan and Beijing. Most cases had a travel history to Wuhan (42%) or where residents of Wuhan (44%), while 14% had no direct relation to Wuhan.

**Table 1:** Characteristics of 2019-nCoVpatients included in the line list. Data are available here ^10^.

| Characteristics | Patients (N=288) |
| --- | --- |
| Age: median (range) in yrs | 49 (2-89) |
| Sex: Male | 62.3% (172) |
| Location |  |
| Mainland China | 68.4% (197) |
| Hubei*, China | 14.2% (41) |
| Shanxi, China | 12.5% (36) |
| Tianjin, China | 7.6% (22) |
| Yunnan, China | 6.6% (19) |
| Beijing, China | 6.2% (18) |
| Outside Mainland China | 31.6% (91) |
| Relation to Wuhan |  |
| Visited Wuhan | 42% (119) |
| Wuhan Resident | 44% (127) |
| None | 14% (41) |
| Disease Outcome: death at time of reporting | 13.5% (39) |

\*Including 32 from Wuhan

The age distribution of cases was skewed towards older age groups with a median of 44 years for cases who were alive or with unknown outcome at the time of reporting (Figure 1A). The median age of 2019-nCoVcases who had died at the time of reporting was 70 years. Few cases (n=7, 2.4%) were under 15 years of age. Adjustment for the age demographics of China confirmed a deficit of cases among children, with a RR below 0.3. in individuals under 15 years (Figure 1).The RR measure pointed at a sharp increase in the likelihood of reported 2019-nCoV infection starting at age 30 yrs.

**Figure 1:**
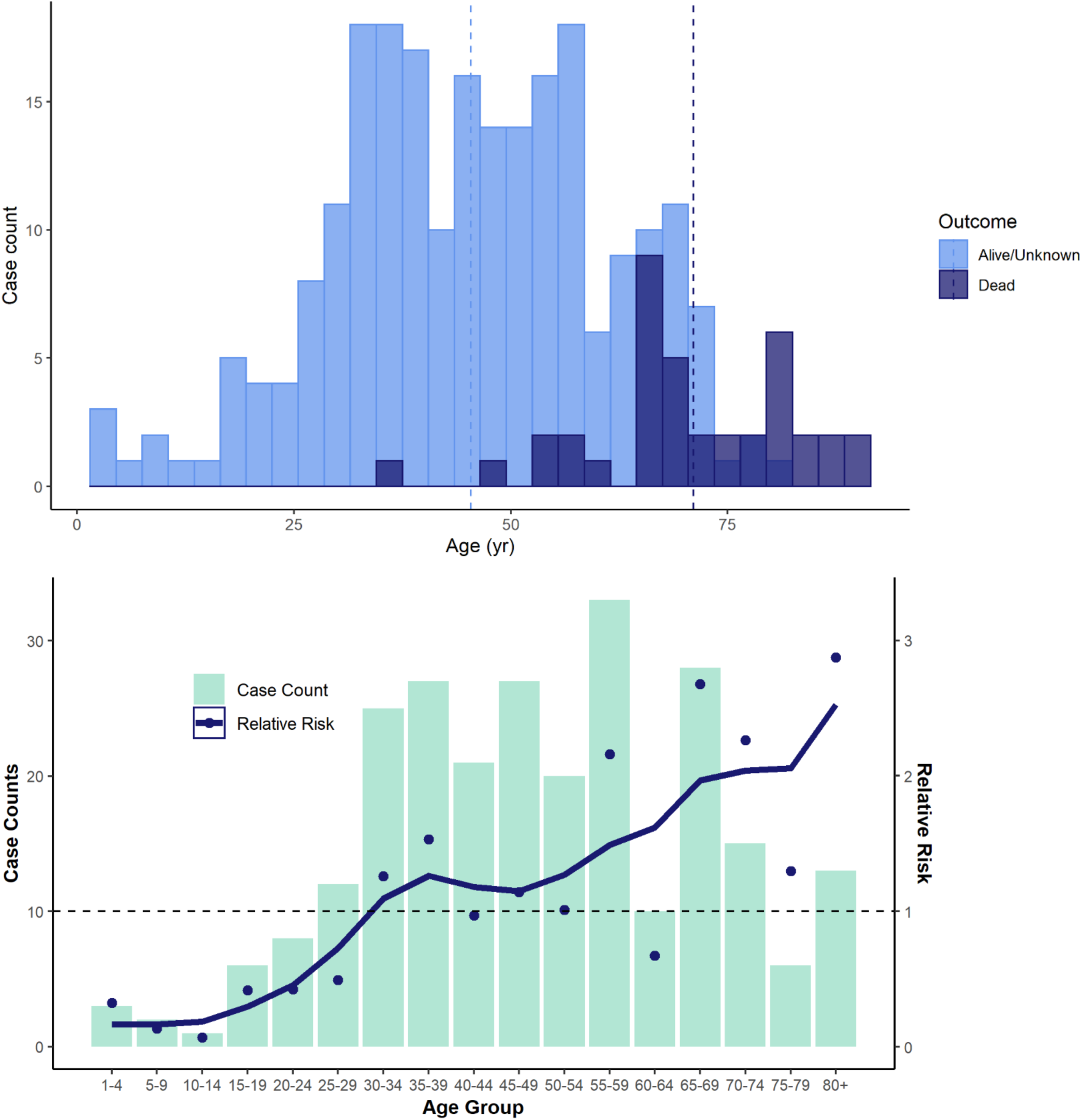
Age distribution of 288 2019-nCoVcases from crowdsourced data. Top: by disease outcome (alive/unknown or dead at time of reporting); vertical bars represent means. Bottom: Relative risk (RR) by 5-year age bands. The observed data is in green and the estimated RR in blue (blue dots=estimated RR, blue curve= spline-smoothed curve)

### Epidemic timelines in Mainland China, nationally and at provincial level

A timeline of cases in our crowdsourced patient line list is shown by date of onset in Figure 2, indicating an acceleration of reported cases by Jan 13, 2020. The outbreak progression based on the crowdsourced patient line list is consistent with the timeline published by China CDC on Jan 29th, based on a more comprehensive database of more than 6,000 2019-nCoVcases (Figure 2). Comparison of cumulative curves shows that the crowdsourced dataset captured a larger fraction of the cases reported to China CDC before Jan 18, 2020 than in more recent days. The very recent slowing down of cumulative cases apparent in the crowdsourced and China CDC curves is likely a reflection of the delay between disease onset and reporting, estimated at a median of 5 days in our data (range 0-43 days).

**Figure 2:**
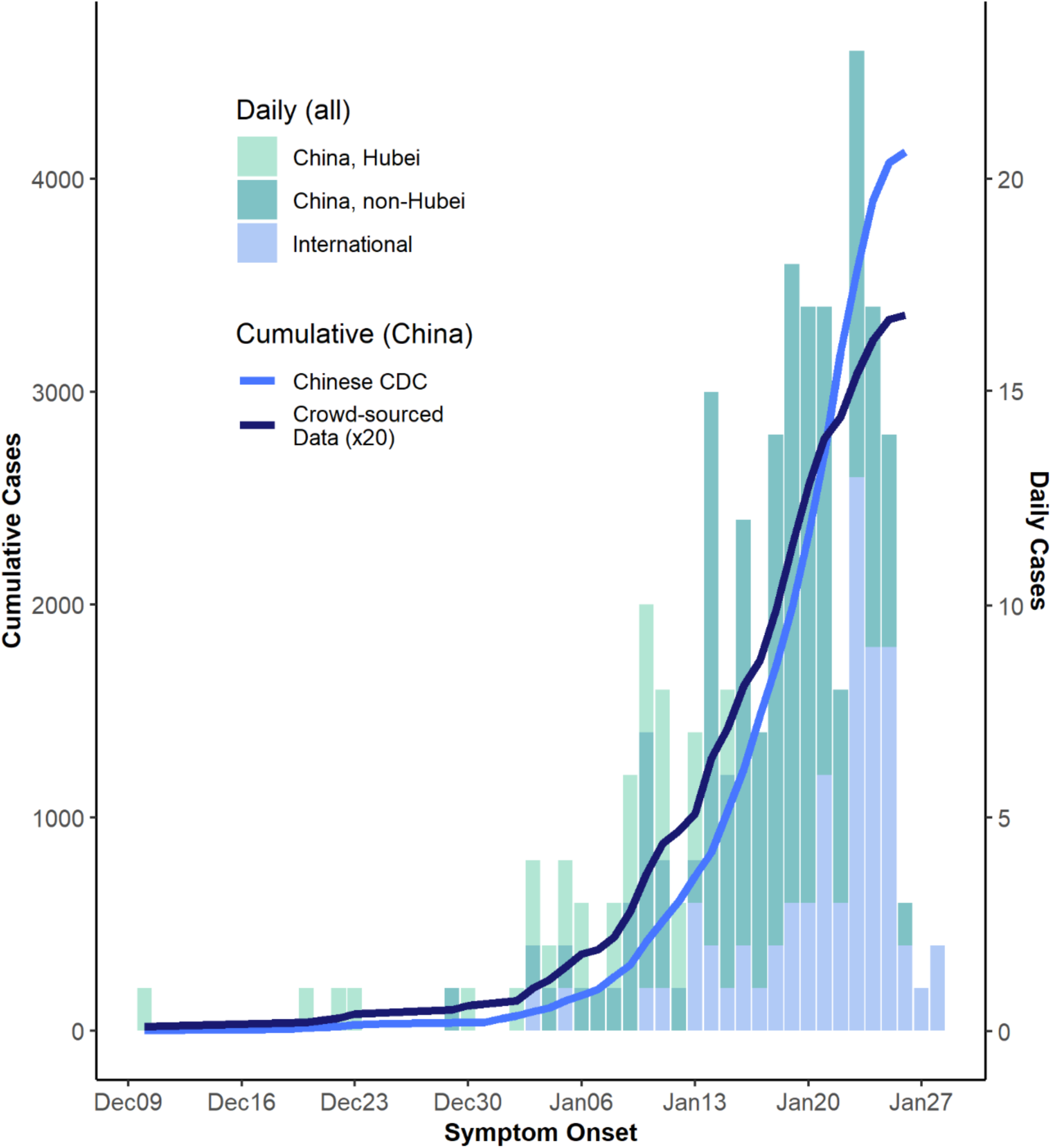
Daily timeline of the 2019-nCoVepidemic based on crowdsourced data and official sources, by location. All data are by date of symptom onset. Cumulative curves are shown for the official China CDC data (first report available on Jan 29^th^, 2020, light blue), and for the crowdsourced data (dark blue). Histograms represent daily case count based on crowdsourced data for Wuhan (light green), mainland China non-Wuhan (darker green), and cases outside of mainland China (light blue).

The subnational growth of the epidemic in China is shown by reporting date in Figure 3. As of Jan 30, 2020, 14 of the 30 provinces had already reported more than 100 confirmed cases. The apparent rapid growth of newly reported cases in many of the provinces outside of Hubei is consistent with sustained local transmission.

**Figure 3:**
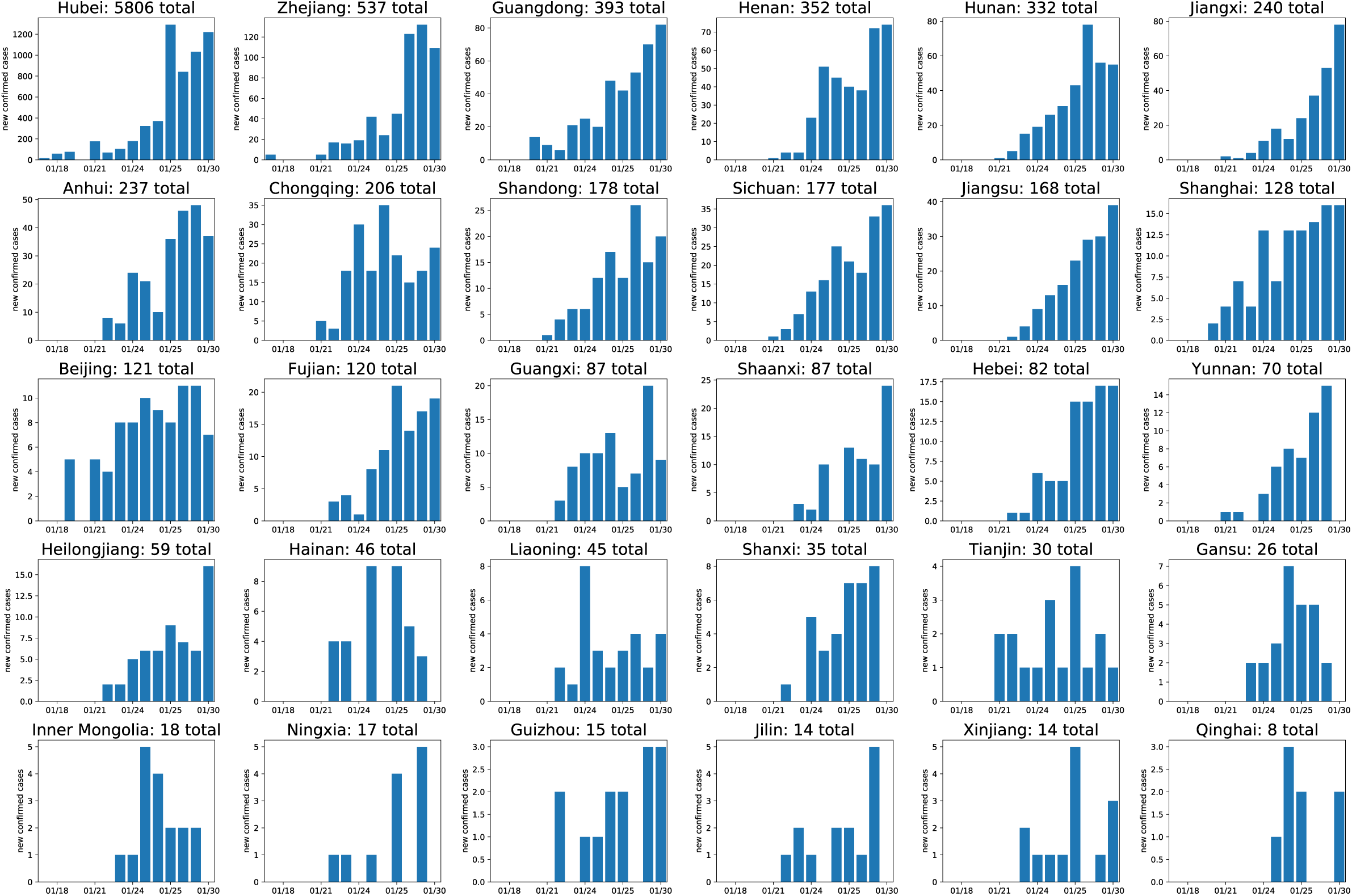
Daily timeline of the nCoV epidemic at provincial level, China, as of 01/30/2020. Curves are by date of reporting and sorted by total number of reported cases. The timeline for each province is reconstructed based on daily outbreak situation reports provided by provincial health authorities and posted on DXY.cn.

### Hospitalization and reporting delays in mainland China

The median delay between symptom onset and seeking care at a hospital or clinic was 3 days in mainland China (IQR 0-15 days) (Figure 4). This delay declined from 5 to 2 days before and after Jan 18, 2020 (Wilcoxon test, p=0.03). Some provinces, such as Tianjin and Yunnan had shorter delays, while the early cases from Wuhan were characterized by longer seeking care delays.

**Figure 4:**
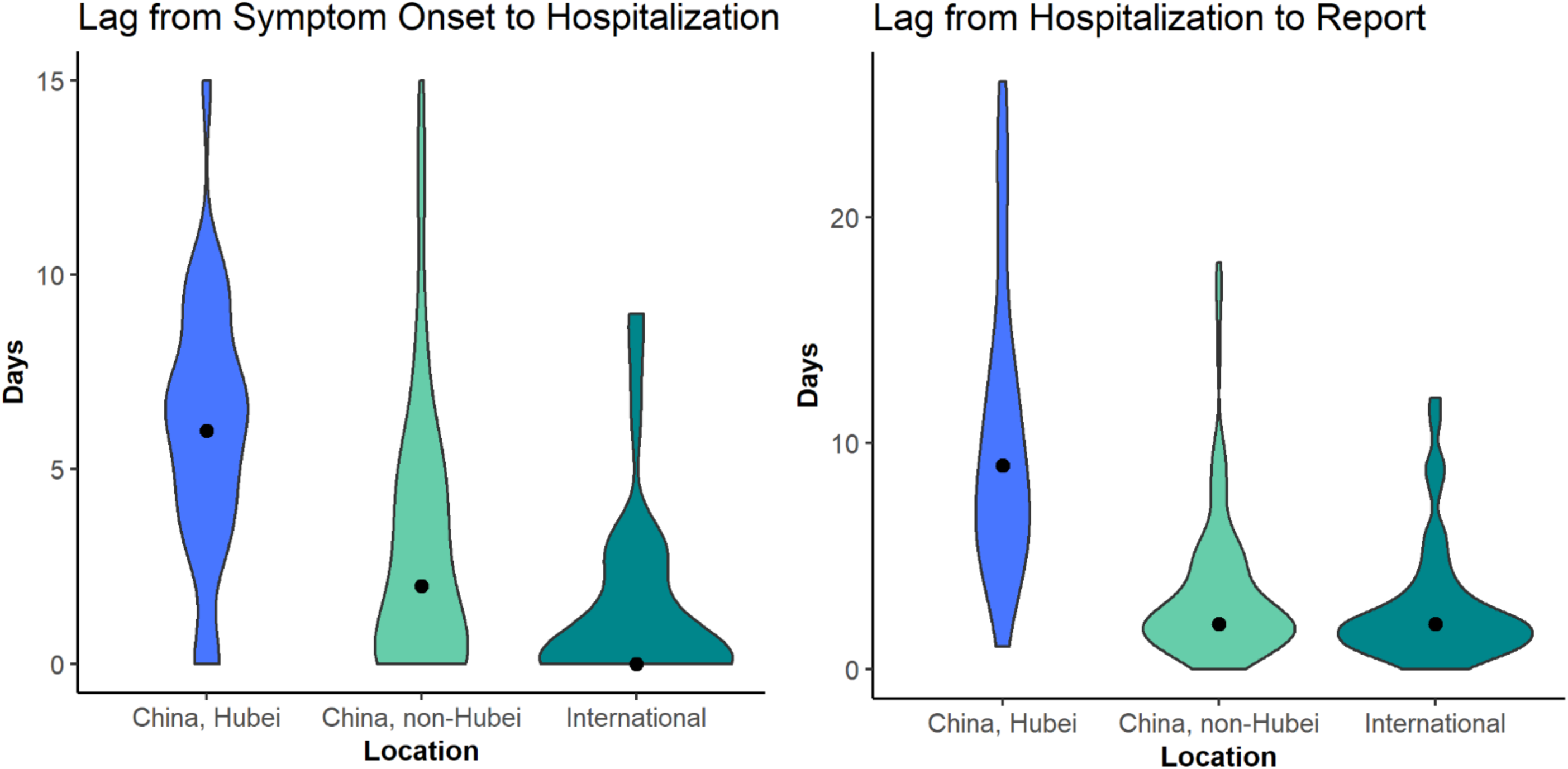
Lags between symptoms onset and hospitalization of 2019-nCoVcases, and between hospitalization and reporting, by location.

The median delay between hospitalization and reporting was 3 days (IQR 2-5 days) in mainland China and declined from 9 to 2 days after Jan 18th, 2020 (Wilcoxon test, P<0.0001, Figure 4). Similar to hospitalization delays, reporting was quickest in Tianjin and Yunnan and slowest in Hubei province.

### Hospitalization and reporting delays for international travelers outside mainland China

The median delay between symptom onset and seeking care at a hospital or clinic was 0 days (range 0-9 days) for international travelers, shorter than for cases in Hubei or the rest of mainland China (Kruskal Wallis test, p<0.0001, Figure 4). Even in the period after Jan 18, 2020, when awareness of the outbreak increased, there was a shorter hospitalization delay for international cases than for those from mainland China (p<0.001). Further, for international cases, the delay between hospitalization and reporting was 2 days (IQR 0-12 days), also shorter than for mainland China (p<0.0001).

## Discussion

Information from patient line-lists is crucial but difficult to obtain at the beginning of an outbreak. Here we have shown that careful compilation of crowdsourced reports curated by a long-standing Chinese clinician network provides a valuable picture of the outbreak in real-time. The outbreak timeline is consistent with aggregated case counts provided by health authorities (for comparison, China CDC published the first epidemic curve by symptom onset on Jan 29, 2020 ^14^). The line list provides information on the lag between symptom onset and detection by the healthcare system, reporting delays, and travel history, which is not available from aggregated case counts published by official sources. Crowdsourced data can contribute to assessment of the effectiveness of interventions and the potential for widespread local transmission beyond the initial foci of infection. In particular, shorter delays between symptoms onset and hospitalization accelerate detection and isolation of cases, effectively shortening the infectious period.

Perhaps the most useful feature of our crowdsourced database was a careful curation of travel histories for patients returning from Wuhan, which, along with dates of symptom onset, allowed for precise estimation of the incubation period ^15,16^. A narrow window of exposure can be defined for a subset of cases who had a short stay in Wuhan, at a time when the epidemic was still localized to Wuhan. Several teams have used ours and other datasets to estimate an incubation period of 5-6 days for 2019-nCoV (95% CI 2-11 dys) ^15-18^. This is a useful parameter to guide isolation and contact tracing; the disease status of a contact should be known with near certainty after a period of observation of 14 days ^15^. Availability of a public dataset enables independent estimation of important epidemiological parameters by several modeling teams, allowing for confirmation and cross-checking at a time when information can be conflicting and noisy.

Another interesting finding in our data is the age distribution of cases. There is a heavy skew of reported 2019-nCoV cases towards older age groups, with a marked deficit of children. This pattern could point at age-related differences in susceptibility to infection, severe outcomes, or behavior. On one hand, a substantial portion of the cases in our database are from travelers, a population which tends to be skewed towards adults (although it does not exclude children). Further, because cases in our dataset were captured by the health system, they are biased towards the more severe spectrum of the disease, especially for cases from Mainland China. Clinical reports have shown that 2019-nCoV severity is associated with the presence of chronic conditions ^18,19^, which are more frequent in older age groups. Nevertheless, we would also expect very young patients to be at risk of severe outcomes and to be reported to the healthcare system, as is seen for other respiratory infections ^20^.

Biological differences could play a role in driving these age profiles. Detailed analysis of one of the early 2019-nCoV clusters suggests that all household members were infected, except for a baby, hinting at true biological differences in the risk of infection driven by age ^21^. It has been speculated that prior immunity from infection with related coronaviruses may protect children from SARS infection ^22^, which could also play a role for 2019-nCoV. In any case, if the age distribution of cases reported here was to be confirmed and the epidemic were to progress regionally and globally, we would expect a rise in respiratory mortality concentrated among ages 30 and above. This mortality pattern would be markedly different from the profile of 2009 influenza pandemic, where excess mortality was concentrated in those under 65 years ^23^.

The age profile of 2019-nCoVcases is reminiscent of SARS infections, for which a deficit of children was also noted ^22,24^. The SARS epidemic was characterized by a concentration of cases among health care workers due to super-spreading events occurring in healthcare facilities ^22^. We do not see a large fraction of health care workers in the data currently available for 2019-nCoV, although one large cluster was reported in Wuhan (not captured in our data).

Based on our data, the timeline of the outbreak indicates rapid growth in several provinces of China, consistent with local transmission outside of Wuhan. At this point, province-level epidemic curves are only available by date of reporting, rather than date of symptoms, which tends to inflate recent case counts if detection has increased. Further, these province-level curves include both returning travelers from Wuhan (importations) and locally acquired cases, which contribute to overestimate the potential for local transmission. It is worth noting that other lines of evidence suggest that local transmission is now well-established outside of Wuhan, as travel increased just before the Chinese New Year and before the ban was implemented in Wuhan ^10,25^. Accordingly, in our line list, we have information on a few local transmission clusters reported outside of Wuhan. For instance, there is evidence of second-generation transmission in Shanxi on Jan 21, 2020. We recognize that our data source is useful and timely but should not replace official statistics. Manual compilation of detailed line lists from media sources is highly time consuming, and it is not sustainable when case counts reach several thousands. Here we provide detailed data on 288 patients when the official case count is over 7000 (as of Jan 30, 2020) ^10^, representing a sample of ∼4% of all reported cases. At the time of this writing, efforts are underway to coordinate compilation of 2019-nCoV cases from on-line sources across various modeling teams. Ultimately, we expect that a line list of patients will be shared by government sources with the global community; however, data cleaning and access issues may take prohibitively long to resolve. For the West African Ebola outbreak, it took 2 years until publication of a line list ^26^. Given the progression of the 2019-nCoV outbreak, such a long delay would be counterproductive.

In conclusion, crowdsourced epidemiological data can be useful to monitor emerging outbreaks, such as 2019-nCoV (see also ^7^ for a related study in the context of Ebola). These efforts can help generate and disseminate detailed information to the scientific community, particularly in the early stages of an outbreak when little else is available, enabling independent estimation of key parameter estimates that affect interventions. Based on our small sample of 2019-nCoV cases, we see an intriguing age distribution, reminiscent of that of SARS, which warrants further epidemiologic and serologic studies. We also report early signs that the response is strengthening in China based on a decrease in case detection time, and rapid management of travel-related cases that are identified internationally. As a caveat, this is an early report of a rapidly evolving situation and the parameters discussed here could change quickly. In the coming weeks, we will continue to monitor the epidemiology of this outbreak using data from news reports and official sources.

## Data Availability

Data presented in the manuscript are made available to the public.

https://www.mobs-lab.org/2019ncov.html

## Notes

### Competing Interest Statement

The authors have declared no competing interest.

### Funding Statement

This work was supported by the in house research program of the Fogarty International Center, National Institutes of Health

